# Neuromodulation for peripheral arterial disease of lower extremities: A 10-year analysis

**DOI:** 10.1101/2023.05.08.23289694

**Authors:** Michael Kretzschmar, Udoka Okaro, Marcus Schwarz, Marco Reining, Thomas Lesser

## Abstract

**Introduction:** Spinal cord stimulation (SCS) or Dorsal root ganglion stimulation (DRG-S) can improve limb salvage, microcirculatory blood flow, and pain relief in patients with peripheral arterial disease (PAD) who are not candidates for revascularization or who have persistent ischemic-related pain after revascularization This retrospective analysis presents 10-year data on the effectiveness and safety outcomes of neuromodulation for PAD at a single center.

**Objective:** This study evaluated the survival and amputation outcome of subjects who received neuromodulation therapy for the management of PAD. Descriptive outcomes such as Walking distance (m), pain intensity (NRS), opioid consumption (MME [morphine milligram equivalents]/d), and self-rated health (EQ-VAS) were also analysed.

**Methods:** This study retrospectively reviews the health data of a single cohort of 51 patients who received an SCS or DRG-S from 2007 to 2022 in a single German center. Survival rate was determined using the Kaplan Meier (KM) curve and major amputation was determined as the amputation of a major limb above the ankle. Patients who received a toe amputation were excluded from the amputation analysis. Two patients were excluded (2 patients died at 3 and 5 months after implantation. Pain, quality of life, walking distance, and opioid usage were assessed before implantation (baseline), 1, 6, and 12 months after implantation, and then annually (during a follow-up visit). Implant-related complications were also documented.)

**Results:** 51 patients (37 men [mean age 68.9± 10.2], 14 women [mean age (68.7 ± 14.6]) underwent SCS (n = 49) or DRG-S (n = 2) implantation due to persistent ischemic pain. The follow-up mean years ± SD is 4.04 ± 2.73 with a range of 1 – 10 years. Atherosclerosis (40.4%) and diabetic vascular disease (48.9%) were the most common pathologies with 48.9% of patients depending on nicotine. At baseline, patients were classified as Rutherford’s Category 3 (n = 23), Category 4 (n = 15), or Category 5 (n = 9). At 24M, 42/47 patients did not require a major amputation following the implant. All patients reported complete pain relief from pain at rest, and 93.3% reported no pain from walking. A total of 75% of patients were able to walk more than 200m and 87% of patients who used opioids at baseline were off this medication at 24 months. Overall, 93% of patients reported an improvement in their overall health assessment. These improved outcomes were sustained through years 3-10 for patients who have reported outcomes.

**Conclusions:** Our single center data supports the efficacy of spinal neuromodulation for improvements in limb salvage, pain relief, mobility, and quality of life. Also, the data show that neuromodulative therapy has a long-term therapeutic effect in patients with chronic limb pain with Rutherford class 3, 4, and 5 peripheral arterial disease (both reconstructable and non-reconstructable).

## Introduction

Peripheral arterial disease (PAD) is most commonly a manifestation of systemic atherosclerosis in which the arterial lumen of the lower extremities becomes progressively occluded by atherosclerotic plaque. Critical limb threatening ischemia (CLTI) is the most severe manifestation of PAD, characterized by ischemic rest pain which progresses to necrosis, limb amputation, and/or patient death^1-4.^ The varied presentations of peripheral vascular disease have led to numerous classification schemes throughout the literature. Rutherford’s chronic limb ischemia classification most resembles Fontaine’s classification, with the addition of objective noninvasive data. The evaluation for any patient with chronic limb pain should include evaluation of the symptoms described in the Rutherford’s classification (Grade I-III, Category 1-6). An important clinical sign for early diagnosis is claudication (Category 1-3). Here, there is not yet a critical threat to the limb. Transition from stage of claudication to critical ischemia (from Category 3 to 4) is particularly significant, as amputation rates and mortality rates increase significantly. Untreated CLTI patients have a 22% annual amputation and mortality rate. The progression of PAD to CLTI ranges from 10%-29% over 5 years^4-6^ with a 1-year amputation rate range of 3.2% - 25%^4,7^ and a 5-year mortality rate of greater than 50%.^7^ Risk factors associated with PAD and CLTI include smoking,^4,8,9^ diabetes mellitus, ^10-14^ hypertension,^15,16^ cardiovascular diseases,^4,17^ chronic kidney disease,^18^ hypercholesterolemia,^19^ and air pollution.^20^ Claudication, defined as pain or tightness in the lower leg due to insufficient blood flow, is the most common and disabling symptom of PAD.^21^ PAD are typically classified as Rutherford categories ^1-6^. The progression of PAD to CLTI imposes other CLTI complications such as ischemic rest pain, tissue loss (ulceration or gangrene), and limb amputation.^22^ CLTI patients have worse quality of life and systemic outcomes (death, myocardial infarction, and stroke), limb outcomes (amputation, wounds, and infections), and functional outcomes (permanent disability and mobility restrictions) than any other group of patients with atherosclerotic cardiovascular disease.^23^

Endovascular interventions,^4^ surgical revascularizations, primary amputation, pain management,^4,24^, or conservative therapies such as smoking cessation,^1,4,8^ walking or supervised exercise^21,25^ are available treatment options. PAD treatment goals include prevention of disabling amputation,^26^ limiting the extent of amputation,^17,26^ reduction in morbidity and mortality,^8,17^ decrease in pain^4^, and improvement in walking distance.^21^ Data from a randomized, prospective, multicenter study show the benefits of a structured exercise program in that walking time and distance improved for PAD subjects who received a treatment combination of optimized medical care and walking exercise. Patients who received both treatments reported more improvements when compared to subjects who received optimized medical care only or optimized medical care plus stent revascularization.^21^ In CLTI patients with ischemic rest pain, arterial revascularization remains one of the main treatments for achieving adequate wound healing and preserving a functional lower extremity^27^ as a successful revascularization improves transcutaneous oxygen (TcPO_2_) 3-4 weeks post procedure. Improvement in cutaneous oxygenation is required for wound healing. Despite successful revascularization, most CLTI patients face a high risk of amputation.^28^ Patients who present late and with tissue loss are most at risk. Amputation rates at 4 years were 12.1%, 35.3%, and 67.3% for Rutherford Categories 4, 5, and 6, respectively.^12^ Additionally, a majority of diabetic patients who are not candidates for revascularization undergo major amputation within 12 months.^29^ Non-revascularization therapies such as prostaglandin treatment,^30^ mesenchymal stromal cells,^31^ wound management,^32^ and neuromodulation therapies such as spinal cord stimulation (SCS)^33-35^ and dorsal root ganglion stimulation (DRG-S)^36^ have been proposed to help patients avoid major amputations and related morbidity.

SCS and DRG-S are alternatives for PAD patients with non-reconstructable CLTI. Multiple studies support the effectiveness of SCS for improvements in limb salvage, pain relief, mobility, lesion/wound healing, and quality of life.^37-42^ Cucuruz et al. (2022)^37^ report that SCS can enable limb salvage in a high percentage of cases and increase mobility due to pain relief. The study retrospectively observed 34 patients in Rutherford stages 4-6 for limb salvage and changes in pain and walking distance. Limb salvage was achieved in 88% of patients. From a preoperative median score of 50m to a 2-year follow-up score of 150m, patients reported increased walking distance and significantly less pain, p=0.001.^37^ A different single-center, retrospective study of 29 patients evaluated the effect of SCS on limb salvage, ulcer closure, and clinical changes of CTLI patients at 1-year follow-up.^22^ Cyreck et al.(2021)^22^ report a limb survival of 97% one-year post-implant with additional improvements in ulcer closure and quality of life. A systematic review by Asimakidou and Matias (2020)^35^ compared the efficacy of SCS in the treatment of non-reconstructable CLTI to conservative treatment in terms of limb salvage, ulcer healing, pain relief, improvement of microcirculatory parameters, and quality of life after 6 months. According to the study, limb salvage and pain relief improved significantly in the SCS group compared with conservative care. SCS patients also required less analgesic medication.

However, long-term evidence of effectiveness is lacking. There is an urgent need to collect and analyze long-term data. Herein, we present a retrospective study examining the long-term outcomes (up to ten years, mean ± standard deviation follow-up of 4.04 ± 2.73) of 51 patients who received spinal neuromodulation (SCS and DRG) for or PAD (Rutherford Categories 3-5).

## Methods

### Patient selection

This retrospective, single-center study enrolled 51 sequential patients diagnosed with PAD-induced ischemic pain referred for consideration of neuromodulation over 15 years (2007-2022). Eligible patients included patients who were not suitable for revascularization; and/or had persistent ischemic-related pain after revascularization. Patients also presented failure of various interventions (including pharmacotherapy) to relieve pain. The Rutherford scale was used to assess severity in PAD and CLTI patients with tissue loss. Included patients were classified as PAD (Category 3) or CLTI (Category 4-5). All patients had angiographic indications. Most patients had already received multiple interventional radiological or vascular surgical treatments. At the time of presentation for spinal neuromodulation, there were no more intervention (surgery, interventional radiology) options. Exclusion criteria included neuropsychiatric disease (dementia), previous spinal surgery at the intended lead placement level, other active implantable devices (e.g., implantable cardioverter defibrillator), severe infection, severe coagulopathy, and CLTI Rutherford 6. Patients who had a life expectancy of less than one year were excluded.

Patients were included after a successful 5-7 days trial period (pain relief ≥ 50%) with an external device. Patients were implanted with an SCS or DRG stimulator at different time points from 2007 through 2022 hence there are varying study entry and exit periods - Figure 1. Clinical charts and operation records were used to obtain patient characteristics such as age, gender, nicotine use, hypertension, diabetes mellitus, and coronary artery disease. The retrospective evaluation of these data was reported to the Ethics Committee of the Medical Association of Thuringia, which had no concerns (registration number 47647/2022/20).

**Figure 1:**
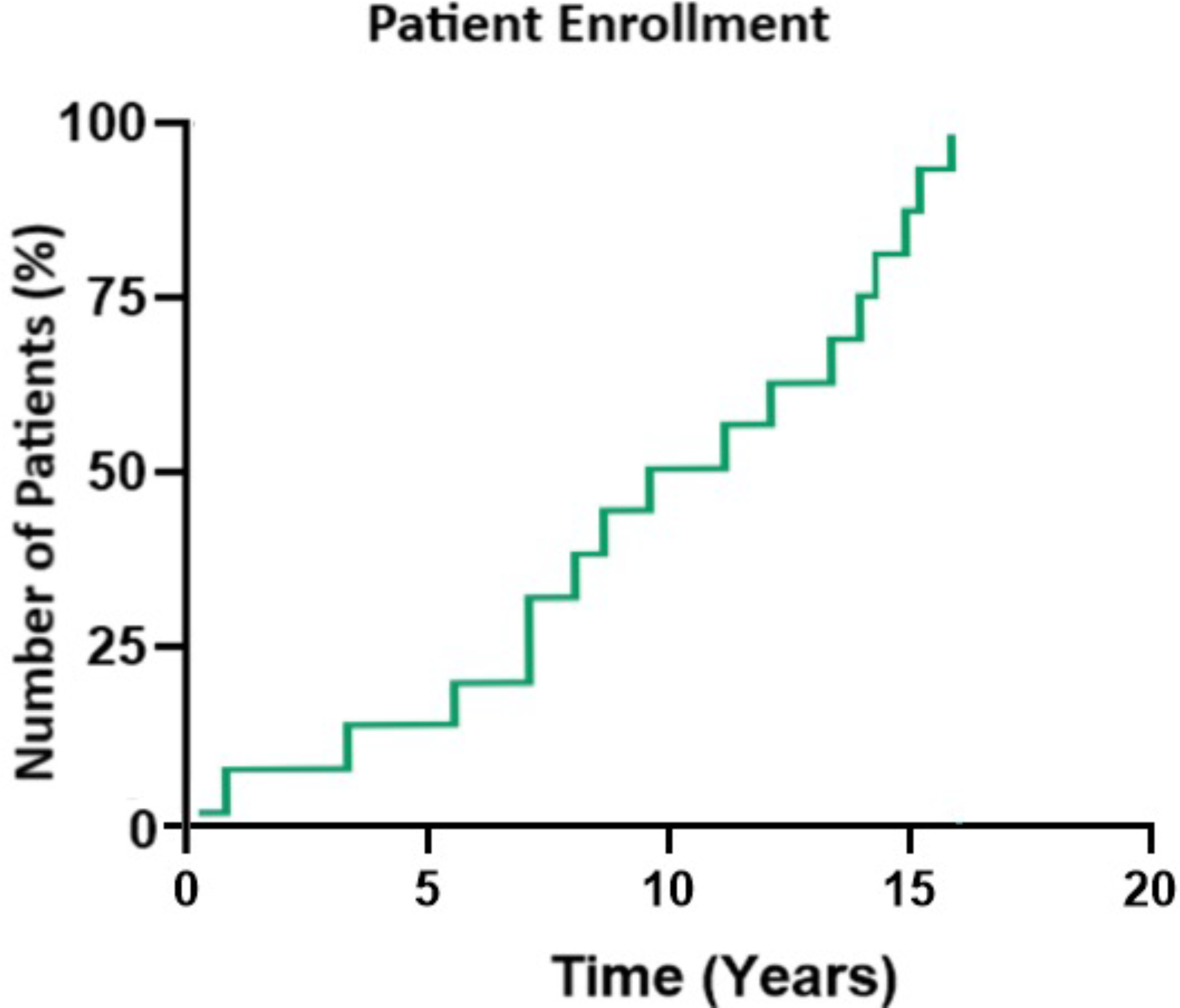
Patient enrollment timeline for this study. The number of patients, N = 51, were implanted at different time points from the first patient implanted in 2007. The Y-axis represents patients’ implant rate. All patients were implanted by 2021.

### Therapy

Enrolled patients were either treated with SCS (n = 49) or Dorsal root ganglion (DRG) stimulation (n = 2). The 49 SCS patients, received either tonic (n = 42) or passive recharge burst (n = 7) stimulation. Patients implanted between 2007 and 2014 received the Eon, Eon-mini, or Genesis implantable generator (IPG) (St Jude Medical/Abbott, Plano TX, USA) with tonic stimulation capability. These IPGs were subsequently upgraded to the Proclaim, ProclaimXR, or Prodigy-MRI IPG (Abbott, Plano TX, USA). Patients implanted from 2015 to 2021 received the Proclaim, ProclaimXR, or Prodigy-MRI IPG which have a paresthesia-free, passive recharge burst stimulation capability. The DRG patients were implanted with the Proclaim DRG IPG (Abbott, Plano TX, USA).

### Procedures

All procedures were performed under monitored anesthesia care. Anxious patients received midazolam (1-3 mg) in combination with sufentanil (5-10 μg) intravenously as a premedication. The operation field was prepared using standard practice of our clinic. Thereafter, local anesthesia using a 1:1 mixture of 0.75% ropivacaine with 1% prilocaine to the operation field was performed. Cylindrical octrodes of (ANS, St. Jude/ Abbott, Plano TX) implanted through a lumbar epidural approach were used for SCS therapy. Test stimulation was then performed to place the electrode (or two electrodes) so that the tingling sensations reached both lower extremities, including the buttocks.

For DRGS cylindric quatrodes (Abbott, Plano, TX) were placed via an epidural approach, with access gained using standard loss-of-resistance technique. Leads were advanced in an anterograde fashion and then directed into the intervertebral foramen near the DRG using curved stylets under fluoroscopic guidance at L4 and L5. Appropriate lead position was determined through intraoperative device programming to confirm paresthesia overlap with the painful regions.

### Study Endpoints

The primary endpoint for this study was to determine the survival and limb amputation rates in CLI patients in response to neurostimulation. The primary endpoint analysis was analyzed using the Kaplan-Meier survival curve on the subjects who received neurostimulation. Limb salvage is defined as the absence of an above ankle or major amputation. Minor amputations such as toe amputation were not included in the limb salvage calculation.

Descriptive endpoints were accessed for pain, walking distance, opioid usage, and patient-reported health assessment. The pain was determined as the reduction of both load-bearing pain (walking pain) and ischemic pain at rest. Reduction of pain was measured using the Numeric Rating Score (NRS). The NRS is a pain scale widely used in medicine to assess pain. The most common form is a horizontal line with an eleven-point numeric range labeled from 0 to 10, with 0 indicating no pain and 10 indicating the most severe pain possible. Walking distance of patients measured in meters (m). Reduction in opioid consumption was measured in milligram morphine equivalents (MME) (mg/day). Overall quality of life was assessed with the EuroQol-visual analogue scales (EQ-VAS) - a patient-reported outcome that asks patients to rate their overall health on a vertical visual analogue scale from “worst possible” to “best possible.”

Endpoint data were assessed at 1, 6 and 12 months after implants, and then yearly for up to 10 years. The data were collected as a standard part of the algesiological diagnostics before the trial, after the implantation and during the follow-up consultation. For the angiological indication of neuromodulation, the walking distance was additionally recorded as an important clinical outcome parameter.

### Statistical analysis

The statistical analysis was performed with Graph pad (GraphPad Software, Dotmatics, Boston, MA, USA). The bootstrapping procedure and tests were performed using SPSS 25 (IBM Corp. Armonk, NY, USA). The Kaplan-Meier survival curve was used for the primary endpoint analysis for survival and amputation rates. T-tests were calculated for descriptive analysis for all pre-implant vs. 1, 6, 12, 24, 36, and 48 months post-implant data for NRS, EQ-VAS, and opioid consumption. Statistical significance was determined if p < 0.05. Because of the observed mortality over the total 10-year period, the 48-month follow-up was chosen as the cut-off time to ensure a sufficient sample size. To compensate for deviations from the requirements for a normal distribution of the analyses, significance tests were based on the bootstrap procedure with 1,000 simulated sample draws each, an alpha level of 5%, and were performed using the T-test for dependent measures. Hedges g, which corrects for smaller sample sizes compared to Cohen’s d, was chosen to represent the effect size. Many consider repeated measures analysis of variance (ANOVA) a standard analytical approach for the given experimental design. However, three reasons justify more adequate statistics. First, an ANOVA tests for any possible difference in outcomes without specific assumptions on when outcomes should deviate from the overall mean or not. Because the effects described here are supposed to reduce negative outcome measures consequently over time (not randomly) and the primary results show very strong differences in dependent measures, an overall test of deviation seems inadequate for this analysis. Second, in case of a significant ANOVA result, further analysis needs to be conducted to determine the exact nature of the difference. Third, the effect size to evaluate the meaningfulness of deviation—eta square—is rather global as well as less specific to interpret. Our data strongly agreed with the predetermined statistical hypothesis. Therefore, it is appropriate to analyze the single reductions or improvements by T-tests for dependent samples and calculate the effect size Hedges’ g from these statistics. Hedges’ g describes the precise differences between baseline and follow-up measures and should be interpreted just like Cohens d, but is additionally correcting for smaller sample sizes. According to the Kolmogorov-Smirnov test, the differences of means of the dependent measures were normal distributed except for a few pain measures. We therefore decided to use a bootstrap procedure based on 1,000 samples for statistical analyses. The diagrams in this publication illustrate the mean values of the dependent measures (NRS, EQ-VAS, and opioid consumption) with corresponding 95% bootstrap confidence intervals. The effect sizes (g) were integrated in the graphs.

## Results

### Patient characteristics

The study enrolled 51 patients (figure 1) - 37 males and 14 females. Two patients who do not yet have 6M data were excluded leaving a total of 49 subjects for this analysis (35 males and 14 females). Patients presented for pain management because of persistent ischemic pain despite exhausted vascular surgery, interventional radiology, and angiologic internal medicine therapy. None of the patients were candidates for revascularization after assessment by the vascular team.

The baseline characteristics of the included patients are presented in Table 1. The mean age ± standard deviation (Age ± SD) of the included population was not significantly different between male and female patients. SCS patients either received the low-rate tonic SCS stimulation (40) or the passive recharge burst stimulation (7) and two patients received Dorsal root ganglion stimulation (DRG-S). The follow-up mean years ± SD is 4.04 ± 2.73. Enrolled patients were classified as CLTI Rutherford Category 3-5 by a vascular surgeon with most subjects experiencing pelvic-leg pain. Diabetes, hypertension, nicotine dependence, and chronic kidney failure were the most common comorbidities and each enrolled patient had at least 2 of the listed comorbidities. Other listed comorbid conditions include obesity and atrial fibrillation.

**Table 1:** Patient Baseline Demographics

| Baseline Characteristics |  |  |  | Patient population (%) | P value |
| --- | --- | --- | --- | --- | --- |
| Sex<br>(Mean Age ± SD) | Male (68.9 ± 10.1)<br>Female (68.2 ± 16.2) | | | 70.2<br>29.8 | P = 0.124 |
| Follow-up Years<br>(Mean years ± SD) | 3.9 ± 2.8 | | | | |
| Neuromodulation Therapy Type | Passive recharge burst<br>DRG-S<br>Tonic |  |  | 14.9<br>4.3<br>80.9 | N/A |
| Rutherford Classification<sup>38</sup><br>(*Grades II and III, categories 4, 5, and 6 are embraced by the term chronic critical ischemia) | **Grade** | **Category** | **Clinical Description** | **No of patients (%)** | N/A |
|  | 0 | Category 0 | Asymptomatic | 0.0 |  |
|  | I | Category 1 | Mild claudication | 0.0 |  |
|  |  | Category 2 | Moderate claudication | 0.0 |  |
|  |  | Category 3 | Severe claudication | 46.9 |  |
|  | II* | Category 4 | Ischemic rest pain | 32.7 |  |
|  | III* | Category 5 | Minor tissue loss-- focal gangrene with diffuse pedal ischemia, nonhealing ulcer, | 20.4 |  |
|  |  | Category 6 | Major tissue loss-- extending above transmetatarsal level, Same as category 5 level, functional foot no longer salvageable | 0.0 |  |
| CLTI Type | Lower Leg<br>Pelvic<br>Pelvic-Leg<br>Thigh |  |  | 2.1<br>18.3<br>59.2<br>20.4 | N/A |
| Medical History | Hypertension |  |  | 76.6 | N/A |
|  | Nicotine dependence |  |  | 48.9 |  |
|  | Type 2 diabetes mellitus |  |  | 46.8 |  |
|  | Occlusion and stenosis of the carotid artery |  |  | 23.4 |  |
|  | Chronic Kidney disease |  |  | 21.3 |  |
|  | Type 1 diabetes mellitus |  |  | 4.3 |  |

### Survival and Amputation Rates

The Kaplan-Meier survival rate for this study was 51.2% and 46.8% at 5 and 10 years (figure 2). A total of 24/49 patients were lost to comorbidities. The cause of death for the deceased patients are listed in Table 2.

**Figure 2:**
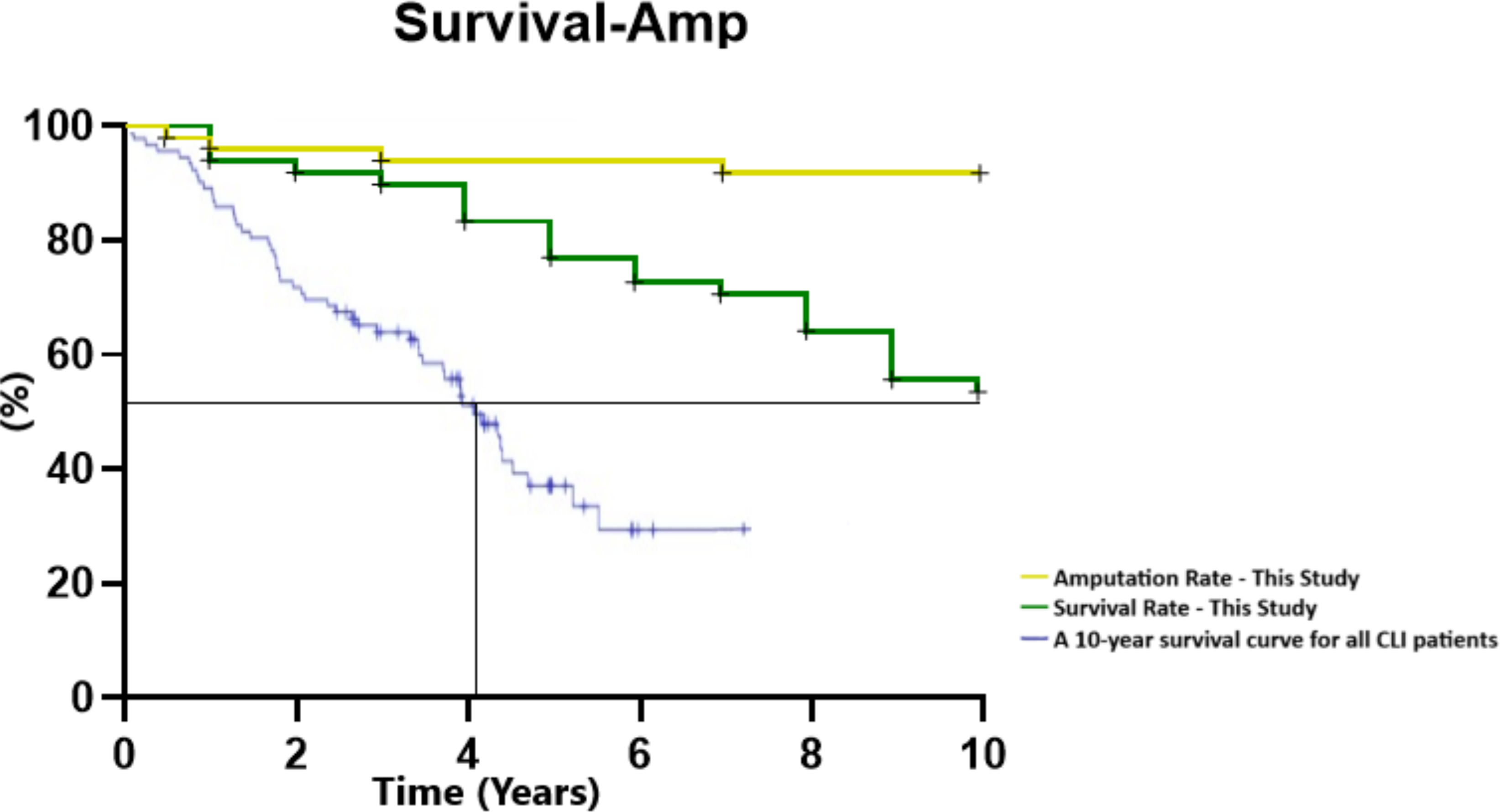
10-year Kaplan Meier survival and amputation curve of all 49 patients. The survival (green) and amputation (yellow) rates are from this study. The comparative survival curve (blue line) is from a similar patient population for both survival and amputation in CLTI patients undergoing treatment (Konijn et al.2020)^39^.

**Table 2:** Reason for Mortality

| **Mortality Reason** | **Number of patients** |
| --- | --- |
| Lung Cancer | 4 |
| Ischemic Heart Failure | 1 |
| Colon Cancer | 2 |
| Acute myocardial infarction<br>(2 patients received amputation) | 4 |
| Chronic lymphocytic leukemia | 1 |
| Glioblastoma | 1 |
| Mesenteric Infarction | 1 |
| Ischemic Stroke | 5 |
| Brain Metastasis | 1 |
| Cerebral Mass Hemorrhage | 1 |
| Left Heart Failure | 3 |
| Sepsis (received amputation) | 1 |
| **Total** | **24** |

The amputation rate in patients undergoing spinal neuromodulation was 10.2% (5 of 49). Five patients (2 Rutherford Category 4 patient and 3 Category 5 patients) were amputated within the study timeframe. One patient received an amputation 5 months after implant, the other 4 received amputation at 8-, 14-, 33-, and 87 months post-implant. A comparison with a similar patient profile study^39^ shows that overall, patients who received an SCS device lived longer with a significantly lower amputation profile. Per Figure 2, the 50% survival rate for the study was reported as 9 years whereas Konijn et al.2020^39^ reported a 50% survival rate at approximately 5.7 years.

Recategorization of patients based on Rutherford claudication symptoms (pain and walking distance), ischemic rest pain, and tissue loss was assessed using the 1-year data. All the patients who presented for the 1-year follow-up were reclassified to category 1 (75.6%, 31/41) for mild claudication, or category 2-3 (24.4%, 10/41) for moderate to severe claudication.

No complications related to the neurostimulation implants were observed in this study. Electrode replacement occurred in 18.4% (9/49) of patients. Implantable generator (IPG) upgrades occurred in 32.7% (15/49) of patients. All IPG upgrades occurred in patients implanted from 2007-2014. These patients received tonic stimulation which was compatible with the IPG.

### NRS Pain Score

Per Figure 3a, patients reported a 91% reduction in NRS score for walking/load-bearing pain (N = 49, Baseline walking NRS score ± SD =7.7 ± 1.1; 24 months score 0.70 ± 1.0, p ≤ 0.0001). Of the 49 patients enrolled, 27 patients presented with resting basal pain. Two were excluded for not having 6M data. The 25 patients were classified as either Rutherford Category 4 (15) or Category 5 (10) patients. The basal pain at rest score ± SD for the 25 patients was 5.3 ±1.5. Patients reported complete pain relief (100%) after 24 months (NRS at 24 months = 0.00, p ≤ 0.0001). Although the number of patients decreases after four years, both pain at rest and pain while walking remains below an NRS score of 2 up to 10 years follow-up (Figure 3b and 3c demonstrate the statistical analysis of the 48-month data). Table S1 reports the average pain score from baseline through 5 years post-implant. Table S2 stratifies the data for patients who reported as Rutherford Category 4 (15) or Category (5) patients.

**Figure 3a:**
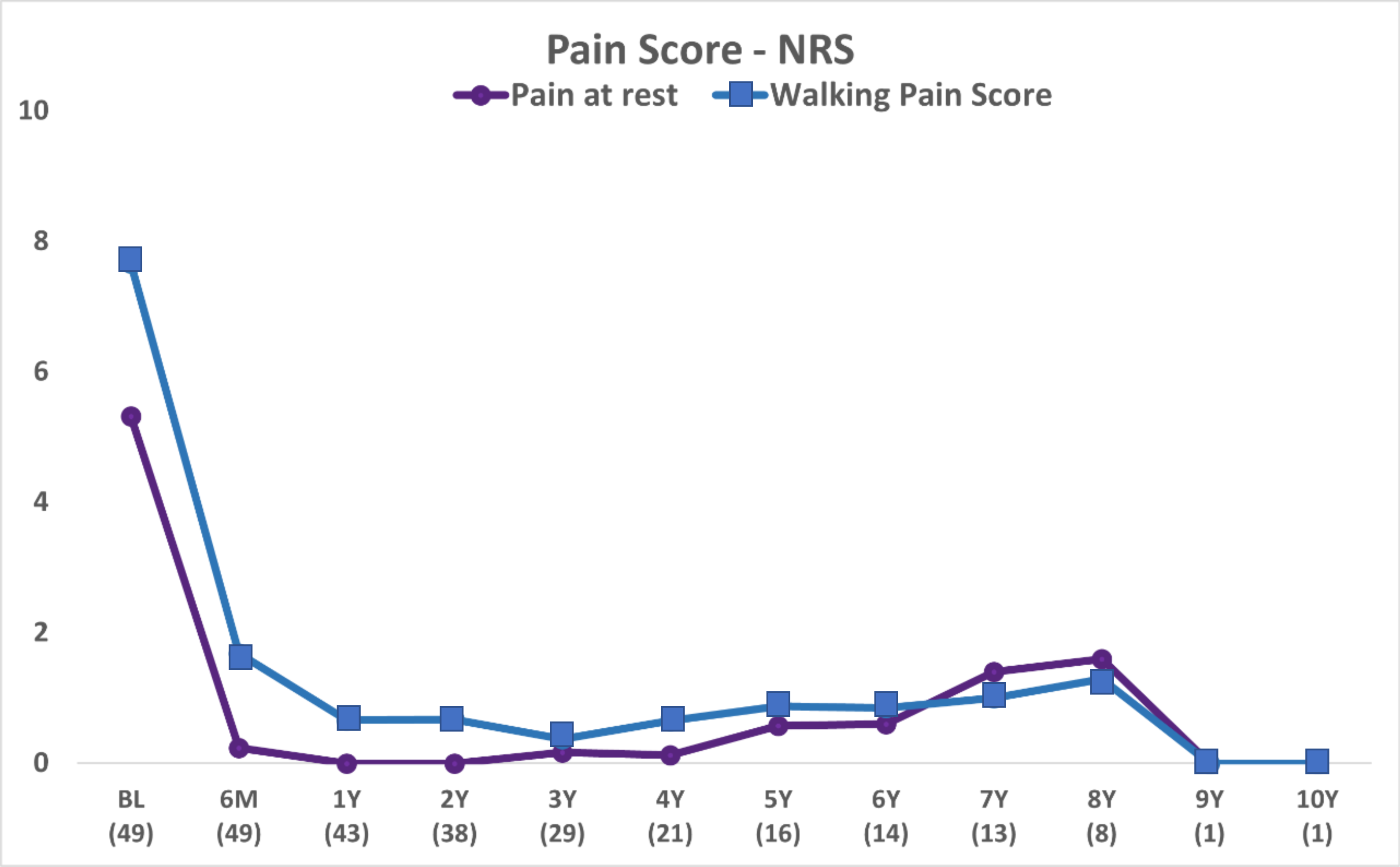
NRS scores for pain at rest and load bearing/walking score. The X-axis presents the follow-up period and the number of patients who present for each visit is included for each follow-up (e.g BL (N = 49). At 24 months, all 38 patients who presented for the follow-up visit reported a 100% decrease in pain at rest and a 91% decrease in walking pain. Arithmetic means are given.

**Figure 3b:**
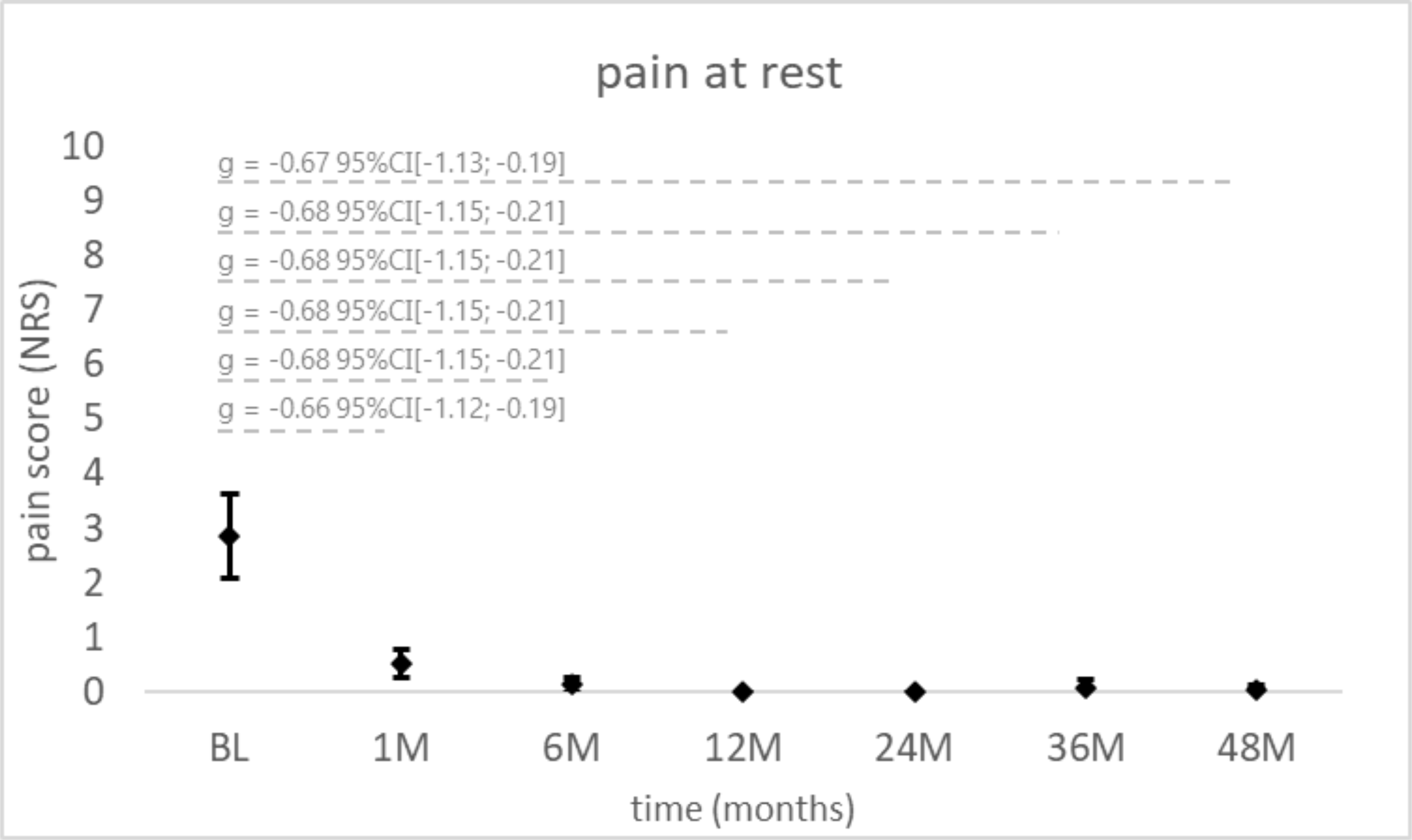
NRS Scores for pain at rest. Descriptive analysis for all pre-implant vs. 1, 6, 12, 24, 36, and 48 months post-implant data. Mean values of the dependent measures with corresponding 95% bootstrap confidence intervals are given, and the effect sizes (g) were integrated in the graph.

**Figure 3c:**
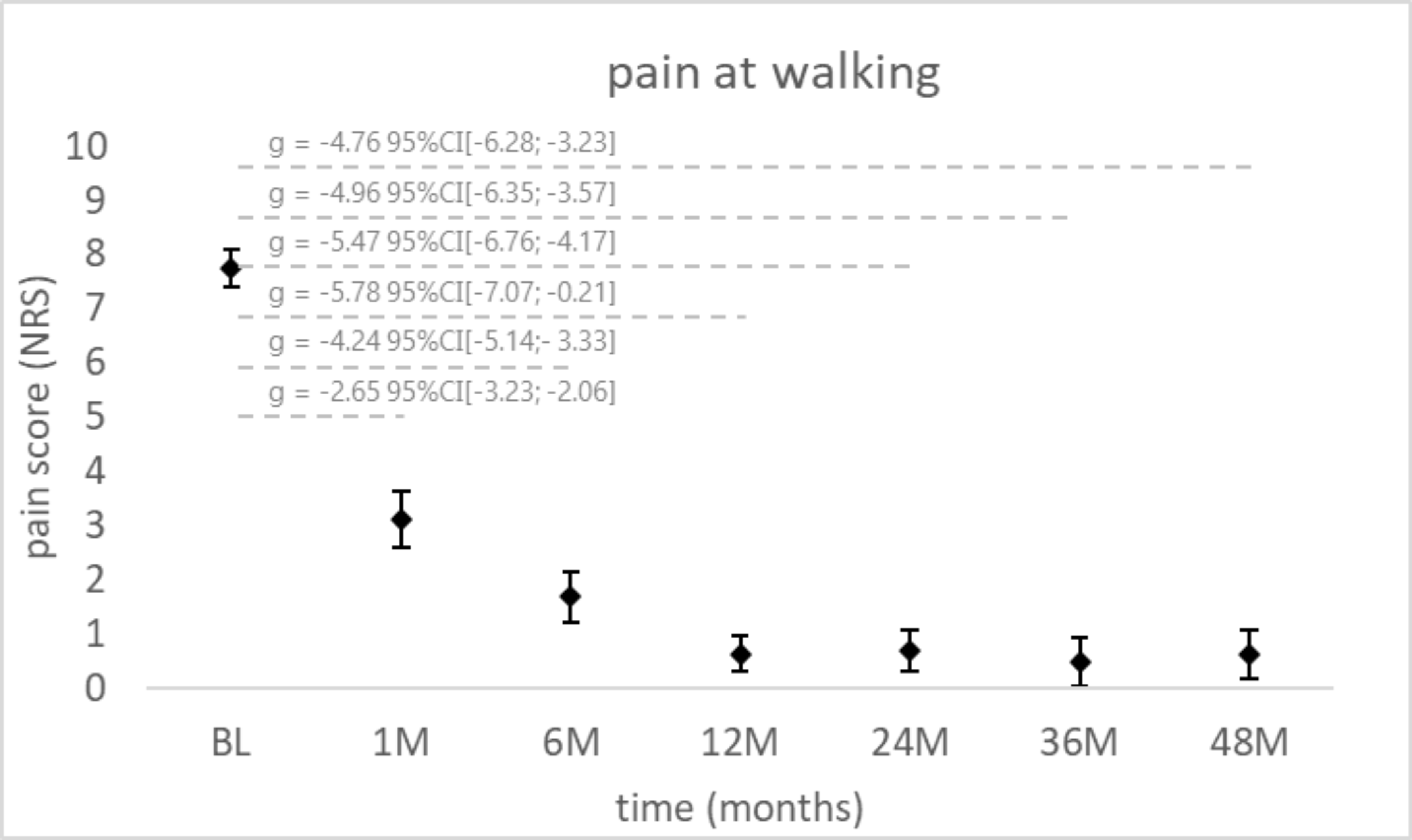
NRS Scores for pain at load. Descriptive analysis for all pre-implant vs. 1, 6, 12, 24, 36, and 48 months post-implant data. Mean values of the dependent measures with corresponding 95% bootstrap confidence intervals are given, the effect sizes (g) were integrated in the graph.

### Walking Distance (m)

At baseline, all 49 patients were unable to walk 200m without severe pain. The average distance (m) ± SD at baseline was 75.5 ± 54.5m. About 71% (27/38) of patients who presented for a 24months follow-up were walking more than 200m without claudication (figure 4).

**Figure 4:**
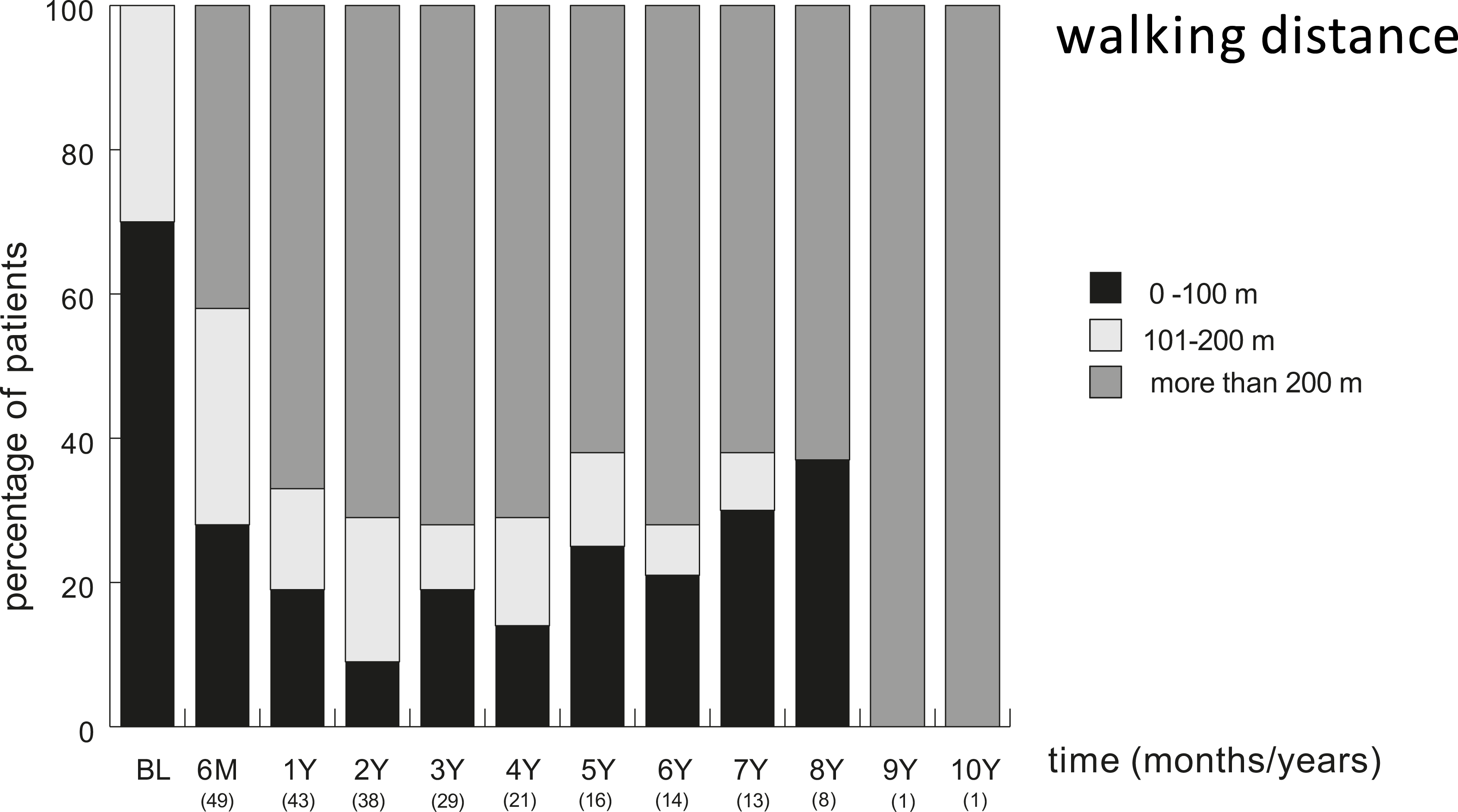
Walking distance (m) for patients. Patients were asked to walk hallways which measure 200m at each visit. The X-axis presents the follow-up period and the number of patients who present for each visit is included for each follow-up (e.g., BL (N = 49). The Y-axis presents percentage of each category.

### Opioid Consumption as MME (mg/day)

A total of 30 patients were on opioid medication (120 ± 60.7 MME, mg/day) at baseline. As demonstrated in Figure 5a, 63% (12/19) of patients reported no opioid usage at 24 months with average use reducing to 22.2 ± 32.0 MME (mg/day). The percentage of patients who were off opioids improved to 80 % of patients at the 48-month visit. Four patients started opioids at years 7-8 but were deceased shortly after. Time of death of these patients is reflected in figure 2 (Years 7-8) and the decline in their health status in Figure 3a (increase in NRS pain scores for Rest and walking at years 7-8) and figure 4 (Decrease in walking distance at the same 7-8 year timepoint). The overall analysis of all patients over 48 months also shows a significant reduction in opioid use (Figure 5b).

**Figure 5a:**
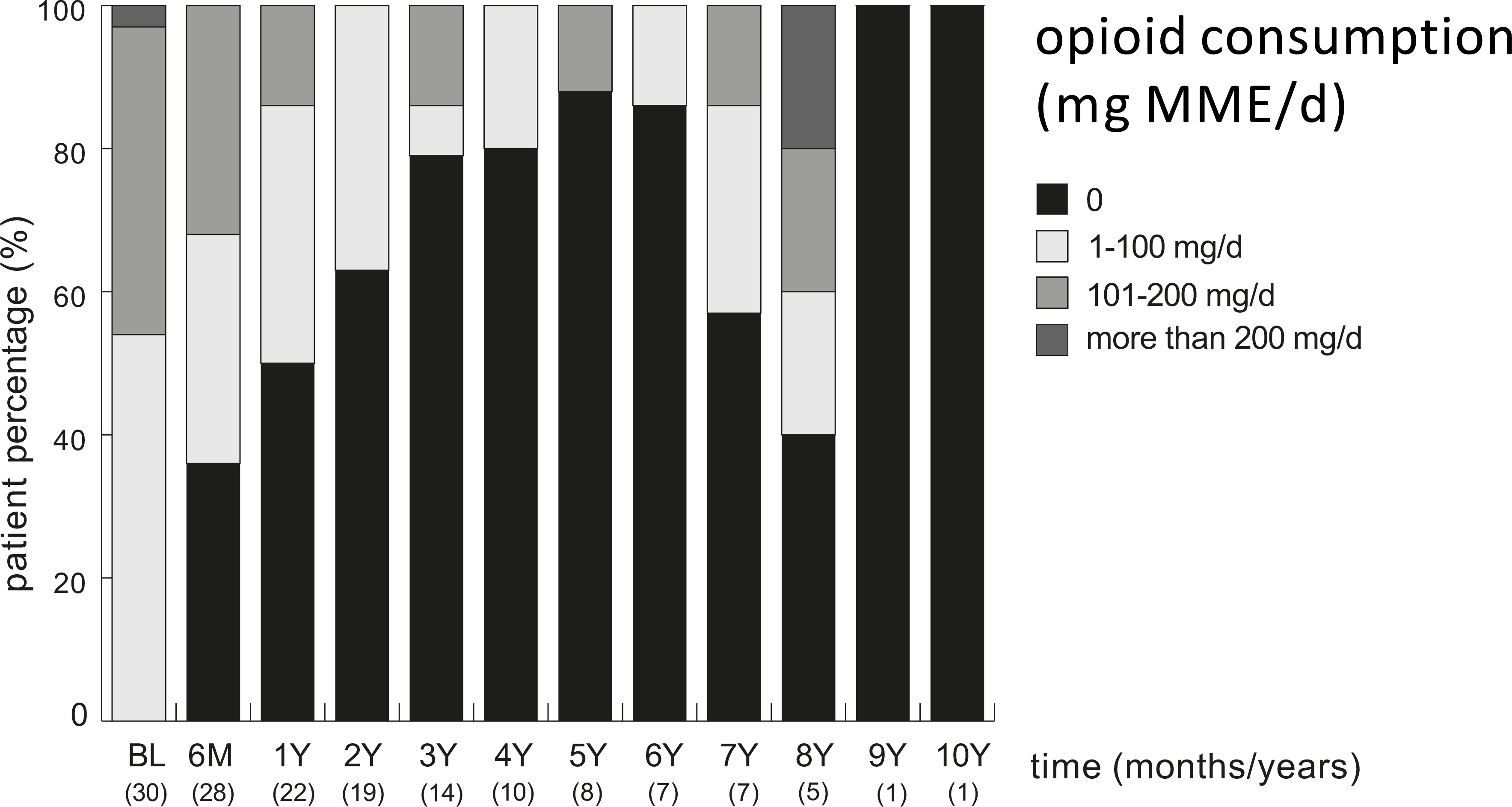
Opioid Consumption (MME, mg/day) for patients. The number of patients who present for each visit is included for each follow-up. Opioid usage was cross-checked using prescription refills and patient reporting. The Y-axis presents percentage of each category.

**Figure 5b:**
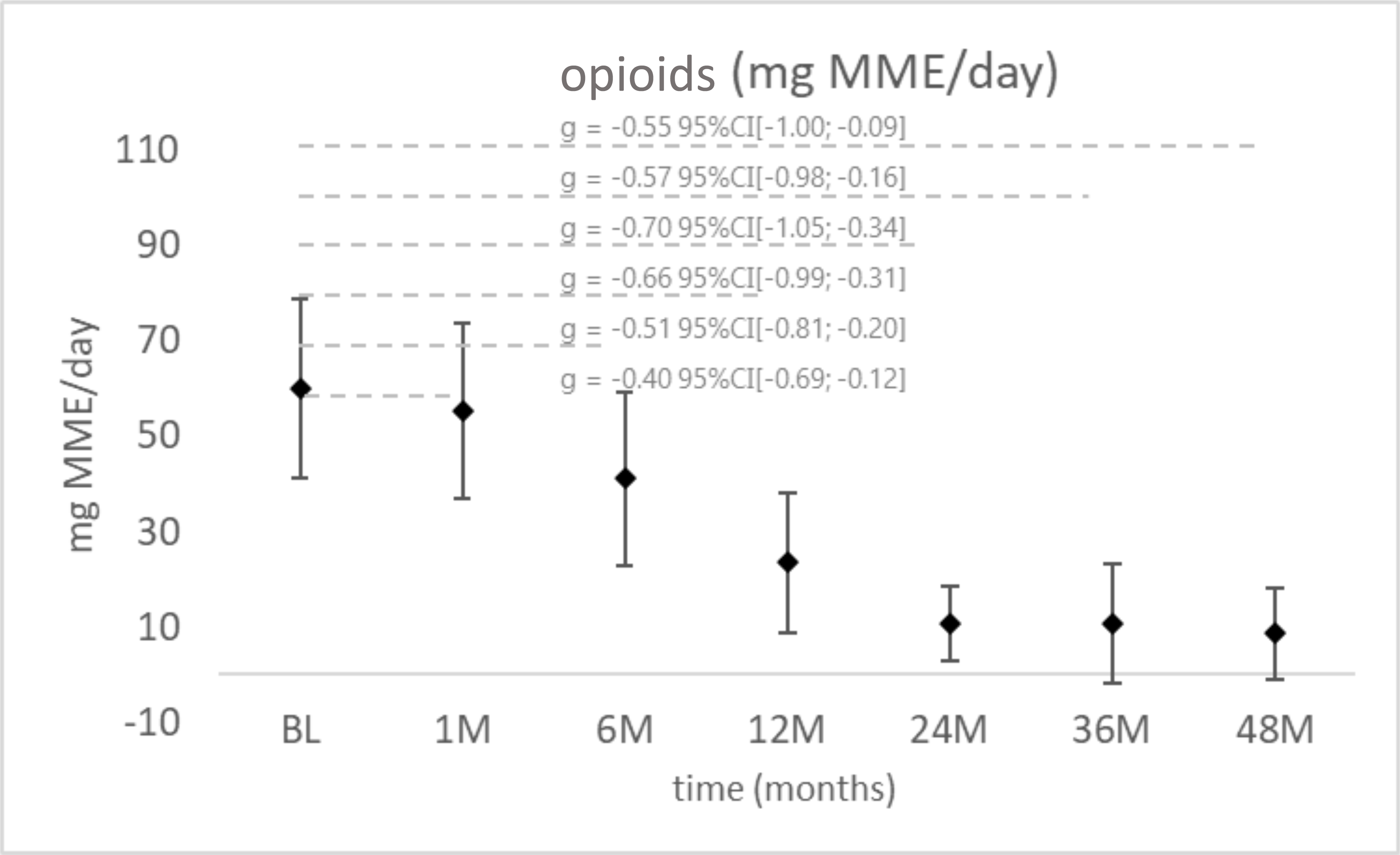
Opioid Consumption (MME, mg/day) for patients. Descriptive analysis for all pre-implant vs. 1, 6, 12, 24, 36, and 48 months post-implant data. Mean values of the dependent measures with corresponding 95% bootstrap confidence intervals are given, the effect sizes (g) were integrated in the graph.

### EQ-VAS Score

All patients’ quality of life improved significantly after the neuromodulation implant. The average ± SD score improved by 44% over 24 months to 65.8 ± 8.4 (baseline score ± SD was 45.6 ± 1 2.1) According to Marten and Greiner (2021)^40^, the average EQ-VAS for a healthy German population is 73.2 (SD = 18.5). (figure 6a-dotted line). When compared to a normal population reference, our SCS patients show a return to normalcy with scores within 1 SD of the population norm. Furthermore, 79% (30/38) of patients reported at least a 10-point improvement in EQ-VAS score after 24 months. This improvement remains stable over the 48-month observation period (figure 6b).

**Figure 6a:**
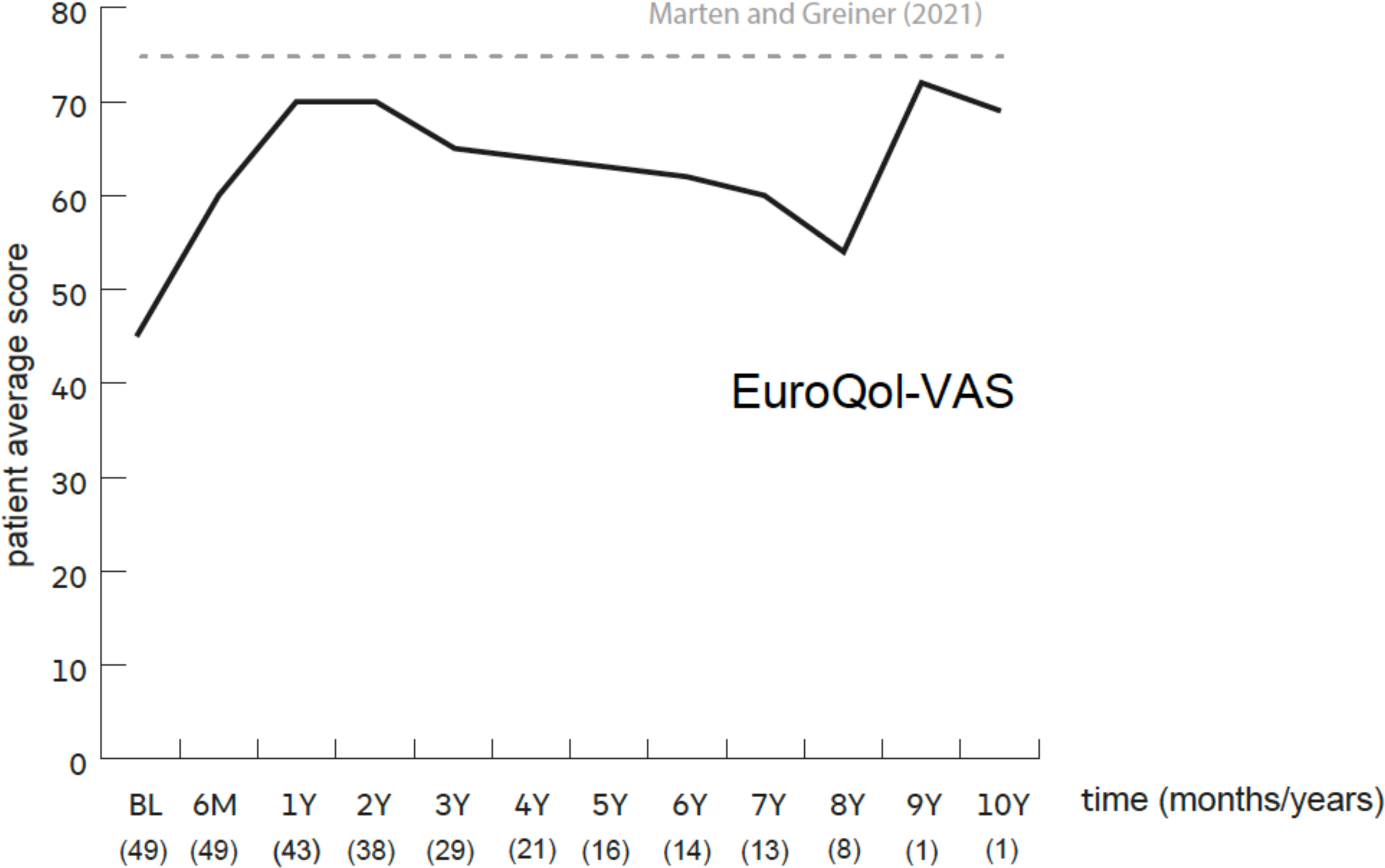
EQ-VAS self-reported by patients. The dotted lines represent the normal population reported by Marten and Greiner (EQ VAS ± SD is-73.2 ± 18.5).^40^ The number of patients who present for each visit is included for each follow-up.

**Figure 6b:**
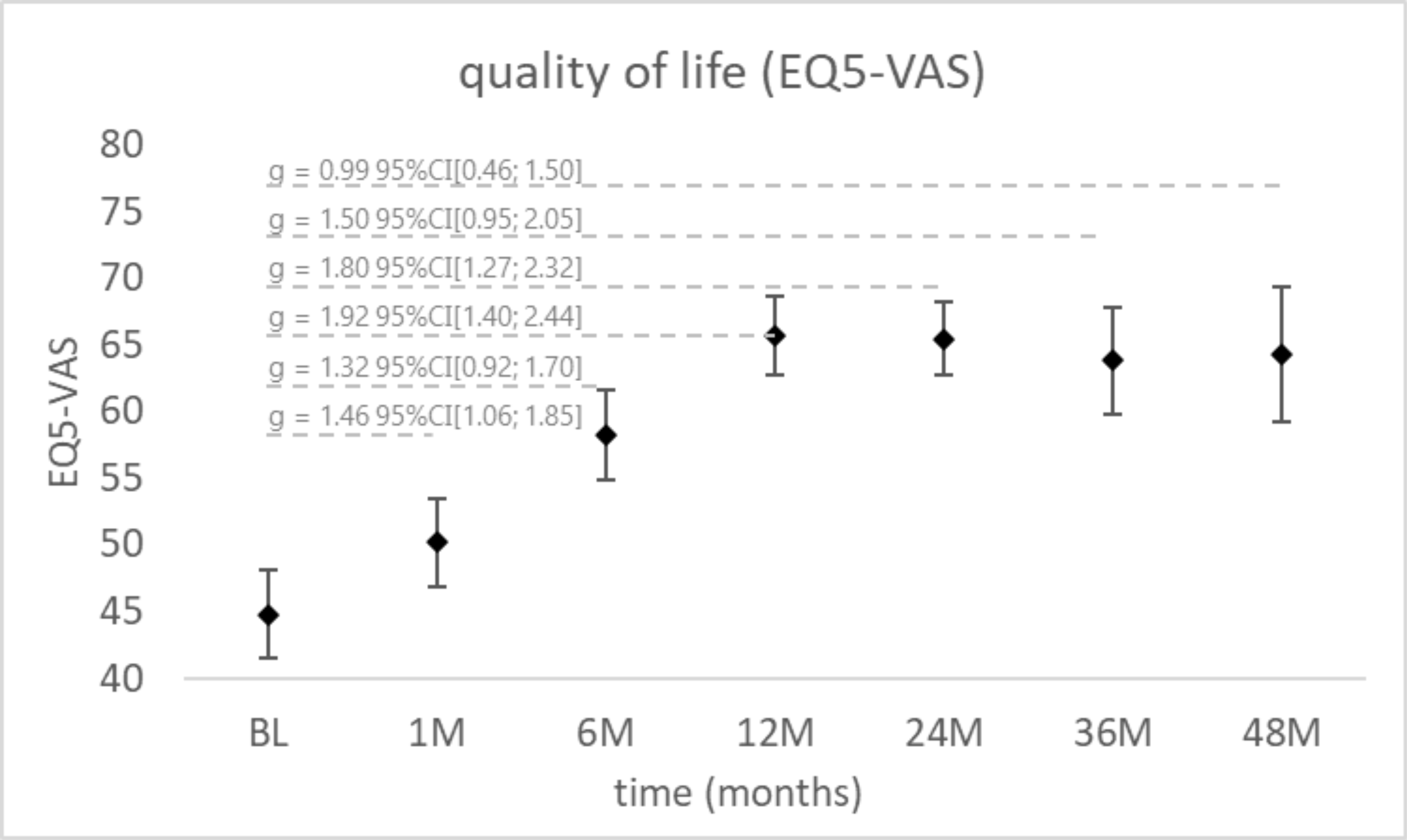
EQ-VAS self-reported by patients. Descriptive analysis for all pre-implant vs. 1, 6, 12, 24, 36, and 48 months post-implant data. Mean values of the dependent measures with corresponding 95% bootstrap confidence intervals are given, the effect sizes (g) were integrated in the graph.

## Discussion

Despite advances in risk factor management and medical therapy, CLTI is associated with significantly increased cardiovascular morbidity and mortality.^4^ A majority of CLTI patients present with co-morbidities such as diabetes^29^ which reduces the limb salvage and survival rates.

Almost half of our patients who received neuromodulative therapy (48.9%) were treated in the claudication stage (Rutherford category 3 = severe claudication). This is not a critical stage which primarily would warrant an intervention/surgery. Nevertheless, from our point of view, the indication for neuromodulative therapy was given because there was no other possibility to relieve the patient of this severe suffering. Therefore, after exhaustion of other possibilities (medication, gait exercise, endovascular or vascular surgery) and after interdisciplinary discussion of the individual case, the indication for neuromodulative therapy was given. This study reports that neurostimulation results in fewer amputations when compared to values reported in the literature; 8.6% for this study compared to 12.1% −67.3% at 24 months.^12,41^ Patients who are not candidates for revascularization observed a reduced survival rate.^41^ In a study of 574 patients with CLTI who did not undergo revascularization after 2 years, 31.6% had died, primarily of CLTI, and 23% required major amputation.^42^ Another study reports the risk of premature death for CLTI patients classified as Rutherford Category 4-6 as 37.7-63.5%.^12^ At the 3-year follow-up, about 25% of the patients were deceased. This study reports a survival rate of 89.4% at 3 years and half of the patients were still alive at 10 years. Nine out of the 23 mortality cases were cancer-related deaths. A comparison with a similar patient population (The Percutaneous transluminal Angioplasty versus Drug-eluting stents for Infrapopliteal lesions (PADI) trial registered in the ClinicalTrials.gov trial register under the number NCT0047) reported a mean survival of 5.4 years.^39^

We report significant improvements in NRS pain scores as well as EQ-VAS, and a reduced opioid consumption. The improvements observed from 6 months post-implant continued to improve through 24-36 months (see Table S1) and were sustained in the patients who presented for a 10-year follow-up. At all-time points, pain intensity (as measured by NRS) was statistically significantly reduced and below an NRS score of 2. The patient reported pain relief 12 months after implant was 100% for patients who originally reported “pain at rest”. This pain relief was sustained across 36 months for the 24 patients who presented with ischemic rest pain. All analyzed patients reported pain during walking and this pain reduced by an average of 90.8% at 24 months. Studies by Ito et al. (2022)^43^ and Klinkova et al. (2019)^44^ both reported 100% pain relief for SCS patients 12-14 months post-implant. Likewise, Chapman et al. (2021)^36^ demonstrated improvements in walking distance, limb salvage, and pain relief. Following the opioid crisis and increasing pressures on healthcare providers to reduce or discontinue long-term opioid therapy, SCS is linked to an increased likelihood of reducing pain medication consumption.^45^ Our study shows a sustained significant reduction in opioid usage. Years 7-8 from Figure 2 - Figure 6 show an increase in pain and opioid consumption accompanied by a reduction in walking and quality of life. These changes were a result of two oncology diagnoses and 1 acute myocardial infarction. All 3 of those patients died shortly after the year 8th follow-up from those diseases.

With the conclusion of the SUNBURST study and approved use of the passive recharge burst waveform in 2017,^46^ the first passive recharge burst patient was implanted in 2017. Of all 7 passive recharge burst patients, there have been no IPG replacements. Waveform comparison shows that passive recharge burst stimulation and classic tonic stimulation produced similar therapeutic outcomes for this study. Patients who received either stimulation reported a range of 22% - 28% increase in EQ-VAS and a 77% - 79% decrease in pain score at 6 months. Two patients who received DRG stimulation reported an 81.3% decrease in pain score at 6 months with an average of 34% increase in EQ-VAS and. The superiority of DRG stimulation to SCS stimulation has been previously published for Complex Regional Pain Syndrome (CRPS) of lower extremity.^46^ Chapman reported superior improvement with DRG stimulation for pain and transcutaneous oxygen measurement (TcPO_2_) - a non-invasive method of measuring the oxygen level of the tissue below the skin^36,47^.

In this study, we observed that the 23 patients who were classified as Rutherford category 3 at baseline, improved significantly more than the 24 patients who were classified as grades II to III at baseline. At 6 months, patients in the former category reported an 89.2% reduction in pain and 73.9% of the patients were walking more than 200m, whereas CLTI patients reported a 68.4% pain reduction and only 16.7% of the population was walking more than 200m. Both SCS and DRG-S have been linked to increases in TcPO_2_ in lower limb ischemia, which improves wound healing and lowers amputation rates.^36,46^ As a result, early neuromodulation intervention may prevent the progression of Rutherford category 3 to CLTI while also improving the patient’s outcome.

Although this study provides additional insight into the potential use of neuromodulative therapy in PAD, there are several limitations. This was a single center study analyzing outcomes from a single provider and approach. Although this may help limit discrepancies in methodology, the patient setting and previous treatments were not standardized. In addition, this was a retrospective report of 51 cases in which data were based on accurate patient information and records. Our limited observations suggest that neuromodulation may have an inhibitory effect on the progression of non contractible PAD (Rutherford category 3). Further studies would be needed to prove this. This would require a prospective study comparing the progression of comparable Rutherford category 3 patients with and without neuromodulative therapy. This therapy could be a valuable option for patients suffering from otherwise untreatable pain. However, it should be noted that only in patients with severe claudication after an intensive evaluation of PAD with imaging and in the absence of revascularization options (e.g., lower leg type, or after unsuccessful revascularization) in an interdisciplinary team (vascular surgery, interventional radiology, angiology, pain management) neuromodulation can ultimately be safely indicated.

Summarizing, this study provides evidence that neurostimulation is effective in terms of sustained pain relief, improved limb salvage, reduced mortality, and quality of life in patients with PAD (Rutherford categories 3-5 and no option for revascularization).

## Conclusions

This 15-year data (2007-2022) effort evidence, that spinal neuromodulation provides a long-term therapeutic effect in patients with non-reconstructable PAD. The data also show that this is a safe form of therapy for these patients, who usually have significant comorbidity.

Neuromodulation reduces pain and amputation rates while improving walking ability and patient quality of life. This is also documented by the reduced need for opioids.

The data may suggest that spinal neuromodulative therapy at the severe claudication stage (Rutherford category 3) delays progression to category 4 or 5. Further studies are necessary to prove this.

## Conflict of Interest

Michael Kretzschmar and Marco Reining were involved in studies organized by Abbott Labs. as Investigators:

(Prodigy I (CRD 694)—A Post-Market Study Evaluating a new Neuromodulation System for the Management of Failed Back Surgery Syndrome or Chronic Intractable Pain of the Low Back and/or Limbs; DELIVERY (CRD 767)—Randomized, ControlleD, Single Blind, ProspEctive, MuLtIcenter Study EValuating Anatomic vErsus TaRgeted Lead Placement for BurstDR TherapY During the Trial Evaluation Period; Prodigy MRI (CRD 800)—A Post-Market Study Evaluating the MR Conditional Neurostimulation Systems).

Marcus A. Schwarz has no conflict of interest to report.

Udoka Okaro is an employee of the Abbott Labs.

Marcus A. Schwarz has no conflict of interest to report.

Thomas Lesser has no conflict of interest to report.

## Data Availability

The data is available in the hospital's own patient data management system.

## Supplementary Data

**Table S1:** Patients’ endpoint calculations over 5 years post implant. P value is generated from a T-test comparison between baseline data and follow-up. Data shows Average (SD), P values are from T-tests for baseline vs. respective timepoint.

| Endpoint | Baseline<br>Average<br>(±SD) | Timepoint Average (±SD) |  |  |  |  |  | P-Value |
| --- | --- | --- | --- | --- | --- | --- | --- | --- |
|  |  | 6M | 12M | 24M | 36M | 48M | 60M |  |
| EQ-VAS* | 45.5<br>(±12.1) | 57.9<br>(±11.5) | 65.5<br>(±9.4) | 65.8<br>(±8.4) | 64.3<br>(±10.1) | 63.8<br>(±12.0) | 62.7<br>(±12.7) | <0.0001 |
| Opioid<br>consumption<br>(MME mg/d)<br>** | 102.9<br>(±60.7) | 69.6<br>(±71.3) | 46.0<br>(±59.2) | 21.2<br>(±32.0) | 11.7<br>(±34.6) | 15.6<br>(±34.3) | 15.0<br>(±42.4) | ≤ 0.01 |
| Pain score<br>basal at rest<br>(NRS)*** | 5.32<br>(±1.5) | 0.21<br>(±0.6) | 0.0<br>(±0.00) | 0.00<br>(±0) | 0.2<br>(±0.6) | 0.1<br>(±0.4) | 0.6<br>(±1.5) | <0.0001 |
| Pain score<br>basal at<br>Walking<br>(NRS)* | 7.7<br>(±1.2) | 1.7<br>(±1.6) | 0.6<br>(±0.92) | 0.70<br>(±1.3) | 0.4<br>(±0.9) | 0.7<br>(±1.1) | 0.9<br>(±1.7) | <0.0001 |

\* = N= 49, Patients who had at least 6M data at baseline
\*\* = N= 30, Patients who were on opioid prescription
\*\*\* = N= 25, Patients who presented with basal pain “at rest”.

**Table S2:** Summary of pain intensity and quality of life data at 12 months according to disease severity. P value is generated from a T-test comparison between baseline data and follow-up. Data shows Average (SD), P values are from T-tests for baseline vs. respective timepoint.

| Endpoint | Patient Population |  |  |  | P-Value |
| --- | --- | --- | --- | --- | --- |
|  | *Rutherford Cat 3 |  | **Rutherford Cat 4-5 |  |  |
|  | 6M | 12M | 6M | 12M |  |
| EQ-VAS | 39.7<br>(±10.6) | 51.9<br>(±12.0) | 61.25<br>(±11.0) | 62.6<br>(±10.0) | <0.0001 |
| Pain score basal<br>at Walking (NRS) | 5.32<br>(±1.5) | 0.21<br>(±0.6) | 0.0<br>(±0.0) | 0.0<br>(±0.0) | <0.0001 |

\* = N= 24, Patients who presented with Rutherford Grade I, Cat 3 at baseline. Patients do not present with pain at rest
\*\* = N= 25, Patients who presented with Rutherford Grade II-III, Cat 4-5 at baseline

